# Voxel-Based Diktiometry - Combining convolutional neural networks with voxel-based analysis and its application in diffusion tensor imaging for Parkinson’s disease

**DOI:** 10.1101/2022.05.25.22275580

**Authors:** Alfonso Estudillo-Romero, Claire Haegelen, Pierre Jannin, John S.H. Baxter

## Abstract

Extracting population-wise information from medical images, specifically in the neurological domain, is crucial to better understanding disease processes and progression. This is frequently done in a whole-brain voxel-wise manner, in which a population of patients and healthy controls are registered to a common co-ordinate space and a statistical test is performed on the distribution of image intensities for each location. Although this method has yielded a number of scientific insights, it is further from clinical applicability as the differences are often small and altogether do not permit for a performant classifier. In this paper, we take the opposite approach of using a performant classifier, specifically a traditional convolutional neural network, and then extracting insights from it which can be applied in a population-wise manner, a method we call *voxel-based diktiometry*. We have applied this method to diffusion tensor imaging (DTI) analysis for Parkinson’s Disease, using the Parkinson’s Progression Markers Initiative (PPMI) database. By using the network sensitivity information, we can decompose what elements of the DTI contribute the most to the network’s performance, drawing conclusions about diffusion biomarkers for Parkinson’s disease that are based on metrics which are not readily expressed in the voxel-wise approach.

## 1. Introduction

Deep convolutional neural networks (CNNs) have become a key element of modern medical image analysis.Traditional versions of the CNNs used for classification involve a series of linear convolutional layers intermixed with non-linear activation and pooling layers. The convolutional layers act as simple image-processing operators, identifying particular features, with the non-linear activation and pooling layers providing both a source of non-linearity at increasingly abstract and coarsely resolved images. At the most abstract level, the features identified may no longer be spatially localised, but encode some information about the content of the image as a whole, which are then used for classification. With the depth and complexity of these networks, it is often difficult to understand and communicate the network’s processing and the “black-box” nature of the network has posed a number of issues for clinical acceptance and integration (Wang et al., 2020).

On the other hand, traditional methods of population-wide whole-brain voxel-based analysis (VBA) such as voxel-based morphometry (VBM) and voxel-based relaxometry (VBR) have become increasingly well-understood and validated in the neuroimaging domain. Arguably, the crux of these voxel-based population-wide analysis is their simplicity: a population is imaged and those images are deformably registered together into some common template space in which the the quantitative intensity (in the case of VBR) or a derived characteristic (such as local scaling in the case of VBM) is used as a univariate distribution upon which one can directly and robustness measure the difference between two sub-populations, normally patients against healthy controls, or to correlate with a different clinically interesting variable often derived from the patient’s symptomatology.

The issue with this approach also arises from its simplicity; it is designed to measure the correlation of a singular area with the clinical variable of interest, not measuring correlations *between* regions that may be of interest. That is, it only identifies regions that simply and strongly correlate with a clinical variable or sub-population, missing regions in which this correlation is weaker or conditioned on some other image feature. The second issue is that these voxel-based methods do not immediately provide a strong prospective method that makes use of their analysis, e.g. classifying new patient into a subpopulation. The individual voxels on their own tend to offer relatively weak classifiers on their own as the statistical analysis only suggests they are better than chance, a relatively low bar for modern classification performance. Recently approaches have used VBA as a method for selecting features to use in machine learning classification, notably support vector machines (Chen et al., 2020)(Prasuhn et al., 2020), which have had variable performance across different disease groups, but illustrate how additional, stronger classifiers would need to be appended to VBA in order to be clinically useful.

Broadly speaking, the general method underlying VBA methods is to use registration and statistical methods to identify potentially discriminative features of a disease, the diagnostic use of these features (including machine learning approaches) is applied afterwards. Depending on how this analysis is performed, larger or smaller regions of interest can be used, but they are pre-specified, rather than determined empirically by classification utility. One possible approach to alleviate these issues is to invert the paradigm by starting with the creation of strong classifiers popularised by deep learning, and use population-wide analysis to understand how the patient images effect the outcome of these classifiers.

The contribution of this paper is to use these techniques for voxel-based population-wide analysis to traditional convolutional neural networks for image classification, specifically the classification between Parkinsonian patients and healthy controls using solely diffusion tensor imaging (DTI). We call this method *voxel-based diktiometry* (VBD) Our goal is to show that traditional CNNs are sensitive to specific characteristic features of diffusion tensors in a non-local manner.

## 2. Theory and Prior Work

### 2.1. Diffusion Tensor Imaging in Parkinson’s Disease

The DTI-based analysis of the brain white matter is a noninvasive imaging approach that has been widely used to measure the diffusivity of the water in the different tissues of the brain, thus allowing the characterisation of the integrity of the tissues associated with normal or abnormal diffusivity and anisotropy values.

In the context of the Parkinson’s disease (PD), VBM, VMR, fixel-based analysis, and tractographic analysis (Cousineau et al., 2017; Li et al., 2020; Xiao et al., 2021) are the more common approaches. Given the dimensionality of the data under analysis, some approaches have opted to perform the analysis on particular regions of interest (ROI), to obtain specific fibre bundles for example (Wasserthal et al., 2019) and some of them have additionally identified the need to also perform correlations between the ROIs (Schuff et al., 2015). Fixel-based analysis follows a similar approach to other VBA methods and thus only a white-matter mask is used to limit the regions under investigation (Li et al., 2020; Xiao et al., 2021).

On the PPMI dataset (Marek et al., 2011) significant alterations between healthy controls and Parkinsonian patients located within the SN, the striatum and the subthalamic nucleus (STN), pallidum, putamen and thalamus have been previously reported (Schuff et al., 2015; Cousineau et al., 2017). Xiao et al. (2021) have also found significant differences in the major white matter bundles between patients and controls specifically on the side of PD onset.

Although the simplest interpretation of PD is that it effects the dopaminergic components of the basal ganglia, the PD is a multisystem disorder involving several other neurotransmitters and pathways (Zhang and Burock, 2020). The distribution of abnormal changes on the DTI values not only at the early stages of the disease but also as possible consequences of neuroplasticity suggest the need to consider a model capable of taking advantage of the heterogeneity of the disease and find these complex correlations on non-predefined neither isolated regions.

### 2.2. Salience in Convolutional Neural Networks

Convolutional neural networks have become a key tool in computer vision. Although originally considered to suffer from the “black-box” problem, where the reasoning of the machine learning tool is difficult or impossible to explain for any given case, CNNs have benefited from a large and early degree of attention towards their visualisation and explanation thanks in part to Simonyan’s “saliency maps” (Simonyan et al., 2013) which led to the development of more advanced methods in explaining the reasoning of convolutional neural networks, sometimes in creative ways, such as with DeepDream (Mordvintsev et al., 2015). These methods rely on propagating the gradient normally used to update the model weights into the input image, either directly visualising it (Simonyan et al., 2013) or using it to modify the underlying image (Mordvintsev et al., 2015). These have the benefit of being simple to implement as well as having an intuitive relationship to the notion of *sensitivity analysis* as the salience maps could be interpreted as the sensitivity of the network output towards a particular region in the image.

## 3. Methods

### 3.1. Patient Images

A total of 213 age-matched individuals (75 F + 138 M, 139 patients (PD) + 74 healthy controls (HC)) with diffusion weighted images were collected from the Parkinson’s Progression Markers Initiative (PPMI) database. The age distribution for the two groups are shown in Fig. 1. The data acquisition was conducted during three to four consecutive years within the majority of the PD patients whereas only during two consecutive years for the HC. The data is comprised of two diffusion-weighting (DWI) samples and one T1-weighted sample per session per year. By assuming a major progression of the disease on the PD group at the last screening, we have selected only the session from the last year for the 139 PD and all the sessions for the 74 HC, thus the total number of samples used for each group is summarized in Table 1.

**Table 1:**
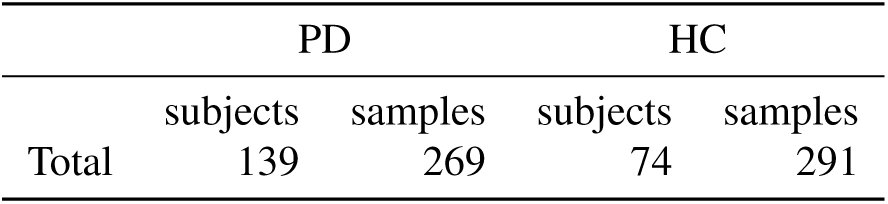
Number of PD and HC individuals and samples.

**Fig. 1:**
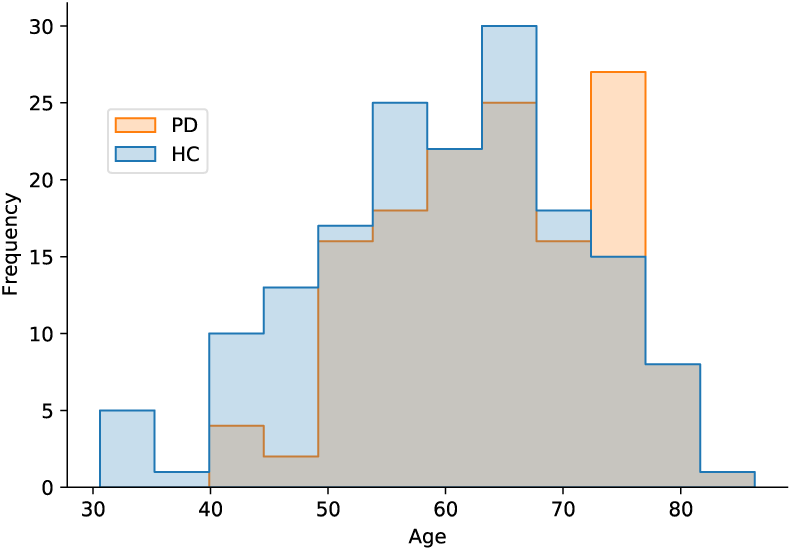
Age frequency distributions of the last screening session for the PD and the two sessions for the HC.

The image acquisition protocol included a 3D magnetization prepared rapid gradient echo (MPRAGE) sequence for mapping anatomical details (TR/TR/TI = 2300/3/900ms; 1 mm isotropic resolution; twofold acceleration; sagittal-oblique angulation) and a cardiac-gated 2D single-shot echo-planar DTI sequence (TE = 88ms, 2 mm isotropic resolution; 72 contiguous slices each 2mm thick, twofold acceleration, axial-oblique aligned along the anterior-posterior commissure) with DWI gradients along 64 sensitization directions and a b-value of 1000s/mm^2^. Repetition time (TR) was in the order of 8,400-8,800ms, depending on the subject’s heart rate (Marek et al., 2011; Schuff et al., 2015). MRI data was downloaded from the PPMI site https://ida.loni.usc.edu/ in DICOM format and converted to NIfTI format using the dcm2niix tool (Li et al., 2016).

### 3.2. Preprocessing

The ROBEX (Iglesias et al., 2011) brain extraction tool was used to extract the brain mask before any other preprocessing step for the T1-weighted (T1w) images. The mask was subsequently used during the co-registration step. Noise removal from the T1w image was performed using the nonlocal means algorithm from the Dipy package (Descoteaux et al., 2008) followed by a bias field correction using the N4BiasFieldCorrection algorithm from the Adavanced Normalization Tools (ANTs) (Tustison et al., 2010). Noise and Gibbs ringing artifacts were removed from the DWI series using dwidenoise and mrdegibbs, respectively. Both tools can be found in the MRtrix3 suite (Tournier et al., 2019). The the eddy_openmp algorithm from FSL (Andersson and Sotiropoulos, 2016) was used to correct for eddy currents and subject movement. The DWI image intensities were then fit to a tensor using the weighted least squares (WLS) method included in SlicerDMRI (Norton et al., 2017).

A deformable registration of the *b*_0_ non DWI with the T1w structural image was calculated using the BRAINSFit tool from 3D Slicer (Johnson et al., 2007) for each subject. The DTI was then resampled in 3D Slicer by preserving the principal direction (PPD) using the transformation previously calculated to be finally co-registered with the T1w in 3D Slicer (Kikinis et al., 2014).

All the images were normalised to have dimension 96 × 112 × 96 (on the sagittal, coronal and axial planes, respectively) before entering to the CNN by adding empty slices or by removing them when needed at each extreme of the volume. Therefore, most of the meaningful information in the centre of the image was retained to serve as the CNN input in a standardised size.

Finally, the images in the patient database were flipped in order to ensure consistent lateralisation of PD to the right side, similar to Xiao et al. (2021). Images from the healthy controls were not flipped (except as a form of data augmentation). This simplifies the network as it only needs to detect PD on the one side, rather than discriminate between the two lateralities of the disease and healthy controls.

### 3.3. Convolutional Neural Network

Since the aim of the proposal is to explore the regions in the image where a CNN focuses its attention, we decided to start this exploratory task by implementing a simple CNN architecture (Fig. 2a). The CNN was trained to classify a DTI into either HC or PD using a supervised approach. The same architecture was used for both the pre-registration and post-registration approaches (see Figure 3b and 3a) in order to keep the model complexity between the two approaches the same.

**Fig. 2:**
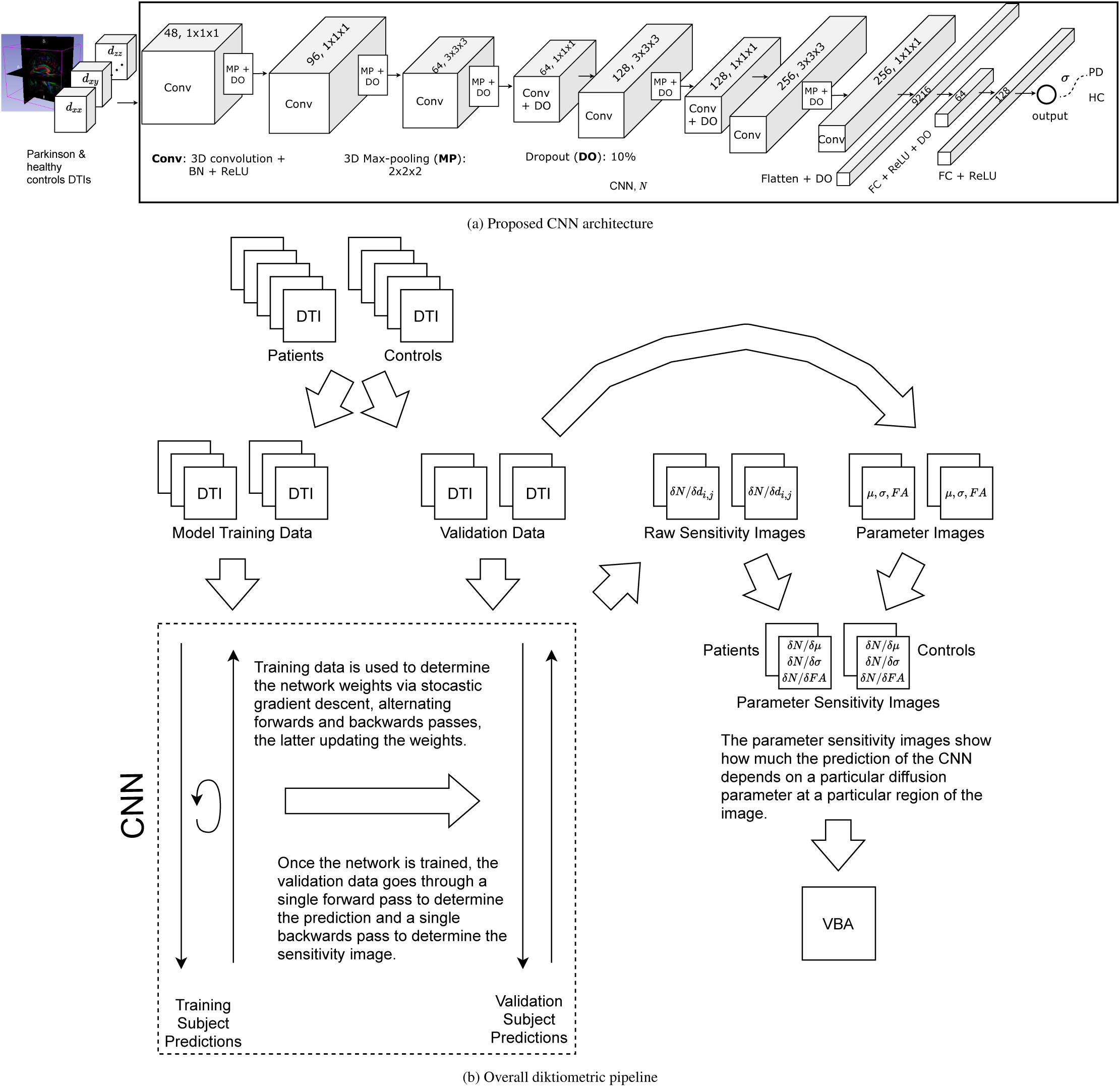
The proposed CNN architecture with 8 convolution and 2 fully connected layers, respectively (a) and, the overall method (b) used to train the CNN and compute the sensitivity images, which are used as the basis for voxel-based analysis.

**Fig. 3:**
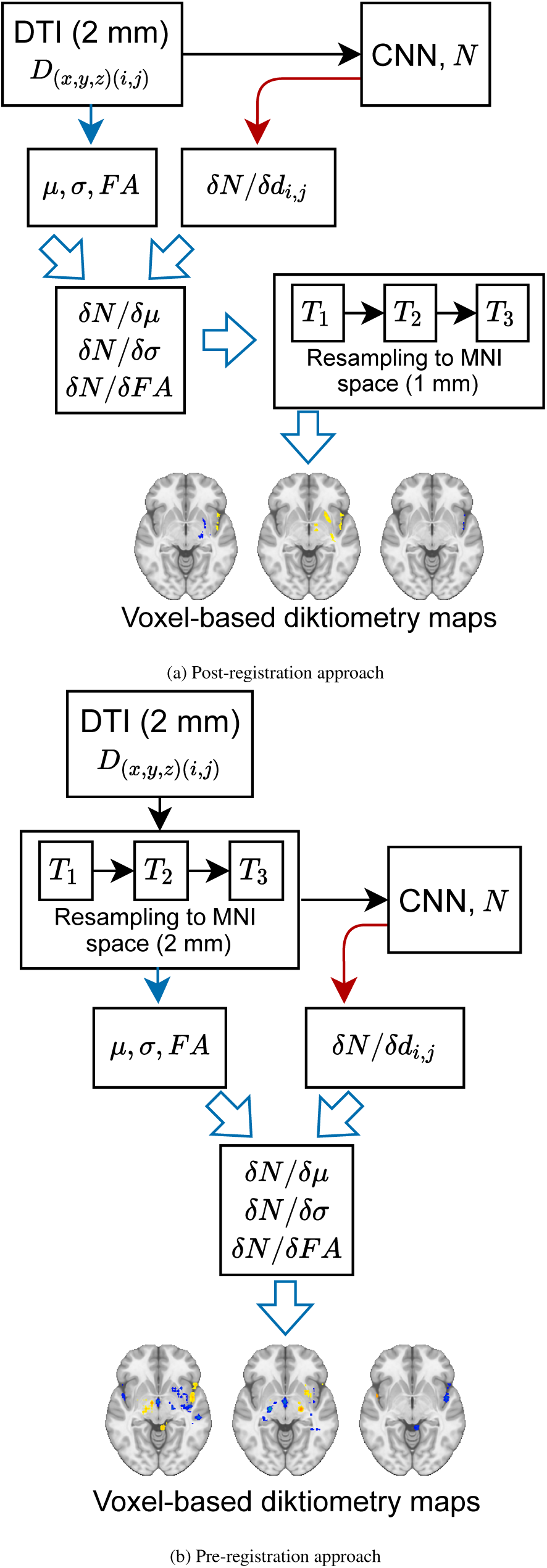
The two approaches to compute the sensitivity images (a) from registered non augmented DTIs and (b) from raw DTIs. The sensitivity images were computed only after the CNN has been trained. The red arrow indicates the backward pass for the gradient computation. VBA is performed over the registered sensitivity images to compute the final VBD maps.

We can consider our network to be some function, *N*(·), that takes an individual DTI, *D*(*p*), as input and outputs the log-likelihood that the image is of a Parkinsonian patient, i.e.

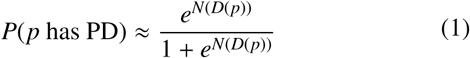

where *D* is the DTI image. Note that increased values of *N*(·) reflects that the network believes the individual is more likely to be Parkinsonian and lower values imply the opposite.

In order to visualise the network’s thought-process, we use a heuristic technique similar to Simonyan et al. (2013) in which the error gradients are propagated through the network also for testing images, not for modifying the network parameters, but for learning how sensitive the neural network is to particular characteristics in particular regions of the image. The overall training and application procedure is given in Fig. 2b.

In order to ensure that the statistical power of later analyses are the same for the diktiometric and the traditional methods, we trained these networks in a 10-fold cross-validation manner which was repeated 10 times. Thus, the sensitivity maps for each individual can be interpreted as arising from an ensemble of ten networks and that each population-wide analysis (described in Section 3.5) is an aggregation of ten networks. The training and testing patients for each fold were randomly selected from the 213 individuals in each repetition. We ensured that all the DTIs from the same individual were either entirely in the training or entirely in the testing subset.

The CNN was implemented in PyTorch and optimised using Adam optimiser with 0.01 L2 regularisation. The learning rate was set to 0.00001 and decreased it every 50 epochs by 4%. We also set a dropout rate of 10% between the convolutional and fully connected layers as shown in Fig. 2a. We trained the CNN for 160 epochs using a batch size of 12.

Random left-right flipping on the HC class was performed during training for both the pre-registered and post-registered versions. For the post-registered version we also added random in-plane rotations up to ±17 degrees and random in-plane translations up to ±10 voxels for both classes.

### 3.4. Tensor shape characteristics

In terms of the tensor shape characteristics, we used the *mean diffusivity*(*μ*), *anisotropy*(*σ*), and *pseudo-planarity*(*θ*):

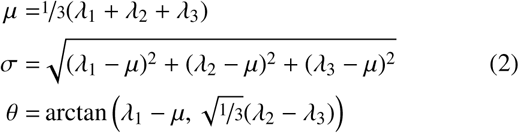

which have the property of having orthogonal sensitivities. The anisotropy in particular is highly related to another, more common metric, the *fractional anisotropy*:

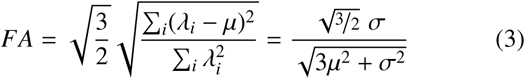

but without the normalisation term in the denominator. This means that the anisotropy’s bounds are [0, 1], but instead 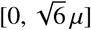. Because of this coupling with both *μ* and *σ*, the fractional anisotropy is not orthogonal and thus it’s sensitivity is coupled to that of the other two metrics:

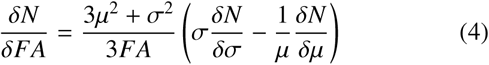

After the image has been processed by the CNN, we performed an eigendecomposition of each voxel in the image to calculate *μ, σ*, and *FA* to create patient-specific diffusion maps. In addition, we computed the sensitivity of the neural network with respect to *μ, σ* and *FA* (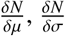 and 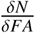 espectively).

### 3.5. Population-wise registration

Unlike in traditional voxel based analysis where a correspondence between voxels in different patients has to be found prior to the construction of the model, there is more flexibility with a CNN-based approach. Registration to find this correspondence can be done either prior to the training of the neural network, providing it with images with a standardised co-ordinate space, or afterwards solely for the population-wise aggregation of the results. The advantage of the first is that the network can learn particular spatially-localised features more readily without having first to detect and localise them. However, this also removes the networks capability of using morphological information, that is, the sizes and shapes of the relevant anatomy have been standardised and their variability is no longer visible. The latter still has access to this morphological information and would be faster for prospective use, as registration would only be necessary for the analysis of a population rather than the individual patient. In addition, it can take advantage of more expressive data augmentation that affects this underlying co-ordinate system (e.g. random shifts and rotations) although it is more difficult to localise particular anatomical features.

The images were all deformably registered to the Parkinson’s disease specific template, ParkMedAtlis (Haegelen et al., 2013), and the MNI template (Fonov et al., 2009, 2011) using the BRAINSFit tool (Johnson et al., 2007).

### 3.6. Voxel-based analysis statistical tests and filtering

The registered parameter or parameter-sensitivity maps can then be used for voxel-based analysis in the common coordinate system. The values for each parameter map at corresponding locations through the patient and control database can then be rigorously compared. These maps were compared using SPM12 (rev. 7771) (Frackowiak et al., 1997) on MATLAB R2014a which assumes a linear relationship between the individual’s status (i.e. PD vs. HC) and the value of the metrics, subject to Gaussian noise. We applied Gaussian smoothing (std. 8mm isotropic) followed by spatial statistical correction with a Family-Wise Error (FWE) rate of 1% and a minimum cluster size of 256 mm^3^.

In order to determine the quality of our approach, we compared it to voxel-based diffusion analysis using the same patient database and registration procedures described in Sections 3.1 and 3.5 respectively.

## 4. Results

### 4.1. Classification accuracy

In order to conclude that the underlying network correctly reflects PD, we first measured the performance of the network and ensured that it is well above that of random chance. Table 2 shows the classification results on the training and testing subsets, respectively. The overall accuracy was on the order of 70% for both the pre- and post-registration approaches

**Table 2:**
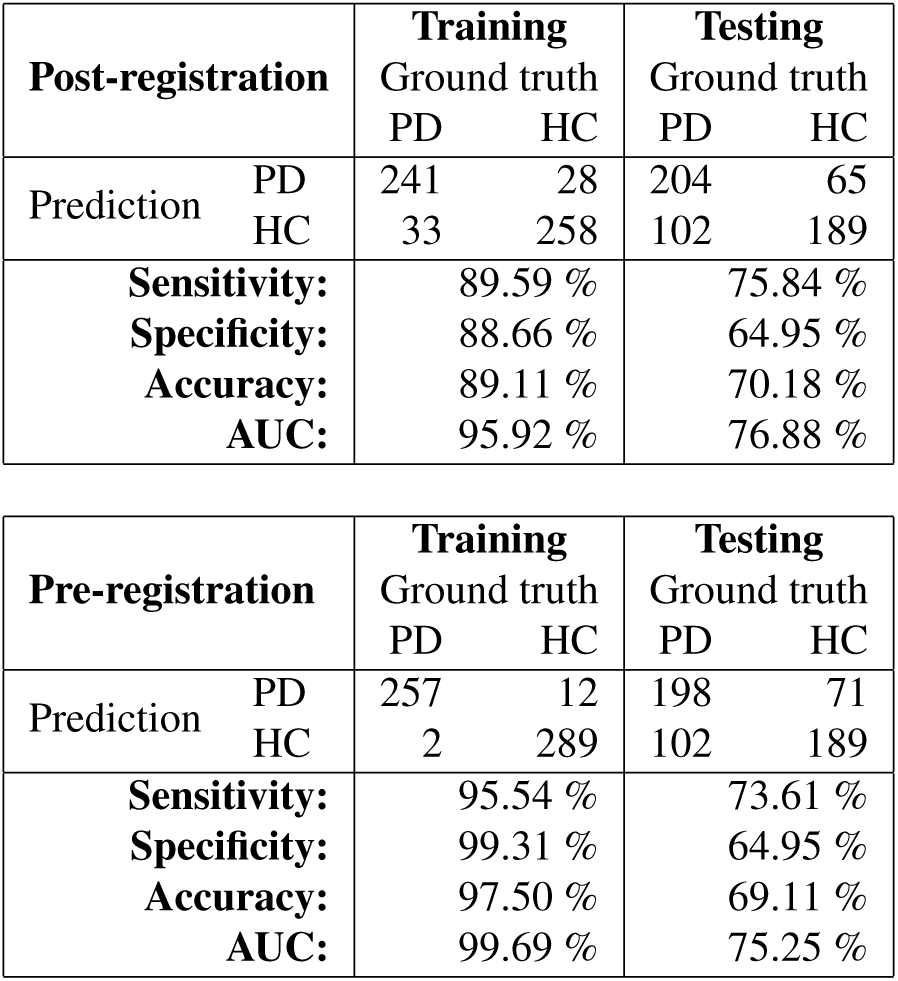
Classification results for the training and testing subsets evaluated on the CNN ensemble.

Regarding the two approaches, we observed a better classification performance and better sensitivity resolution by directly training the CNN from the non-registered DTI images than by registering them in advance. One possible reason could be the loss of morphological information due to the deformation that the structural features suffered when being accommodated into the MNI space from the subjects’ native space.

### 4.2. Voxel-based diktiometry maps

Figures 4, 5, and 6 show the qualitative results for our voxel-based diktiography approach and the comparative voxel-based diffusion approach using SPM and linear statistical analysis and a significance threshold of *p* < 0.01 FWE for the MD, A, and FA metrics respectively. The majority of the results appear in the MD and A metrics for the CNN-based methods, and in the MD and FA methods for the traditional approach.

**Fig. 4:**
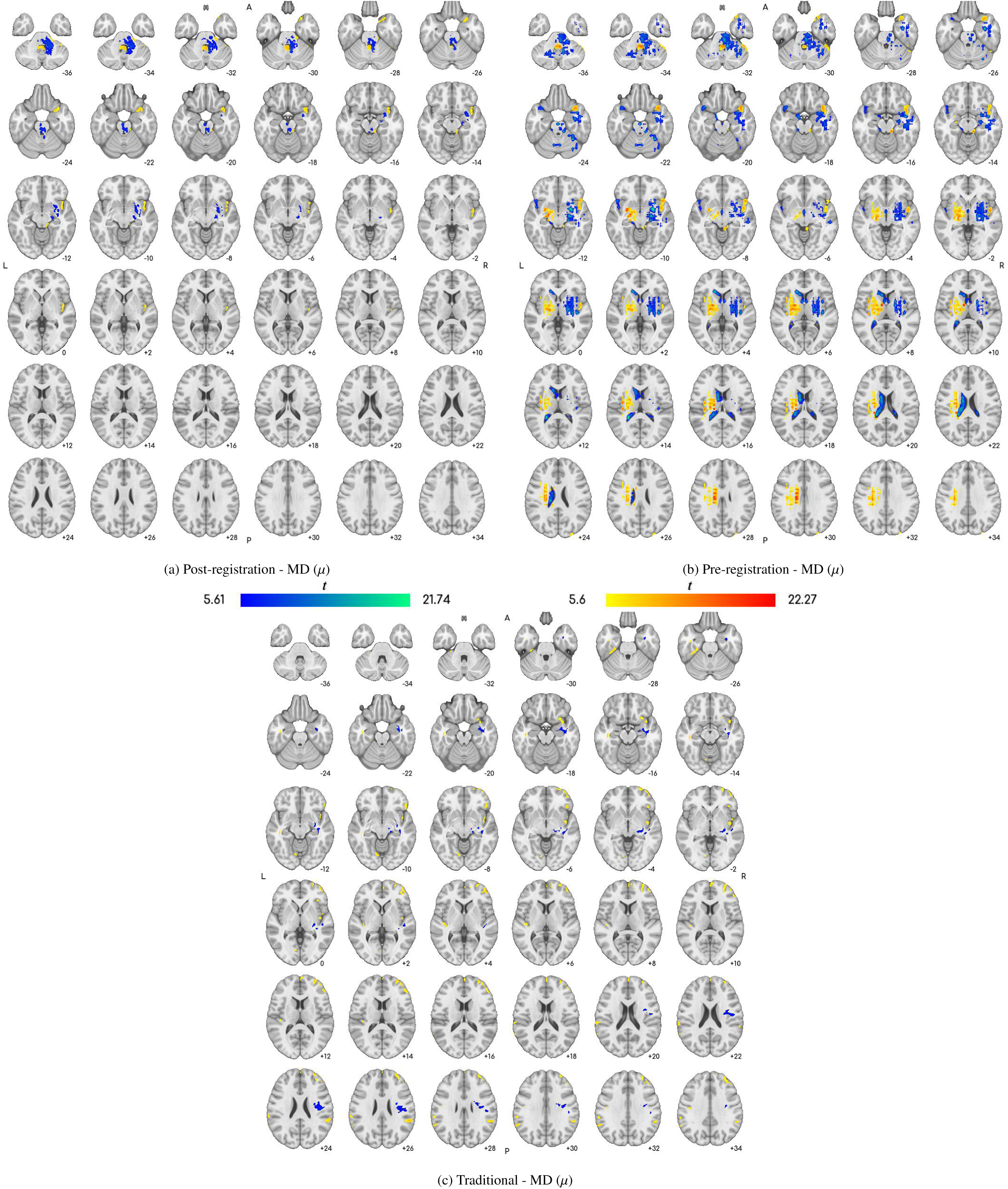
Voxel-based diktiometry (post-registration and pre-registration in the first and second columns, respectively) sensitivity results and traditional voxel-based diffusion analysis results (third column) from *z* = −36 to *z* = +34 in MNI space for the mean diffusivity (MD). Left-dominant PD subjects have been laterally flipped. (Note that patient-right is shown on image-right, i.e. neurological convention.)

**Fig. 5:**
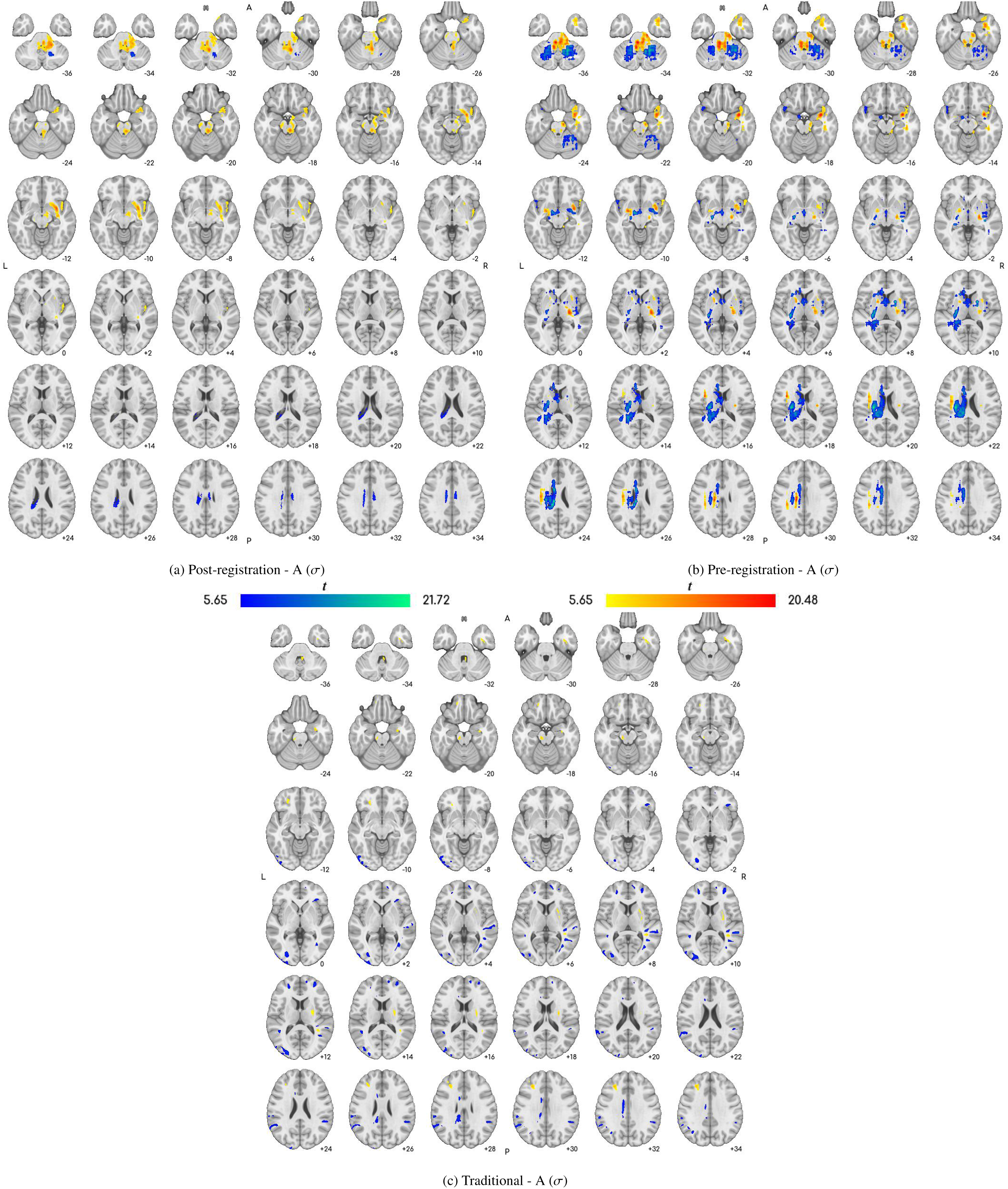
Voxel-based diktiometry (post-registration and pre-registration in the first and second columns, respectively) sensitivity results and traditional voxel-based diffusion analysis results (third column) from *z* = −36 to *z* = +34 in MNI space for the anisotropy (A). Left-dominant PD subjects have been laterally flipped. (Note that patient-right is shown on image-right, i.e. neurological convention.)

**Fig. 6:**
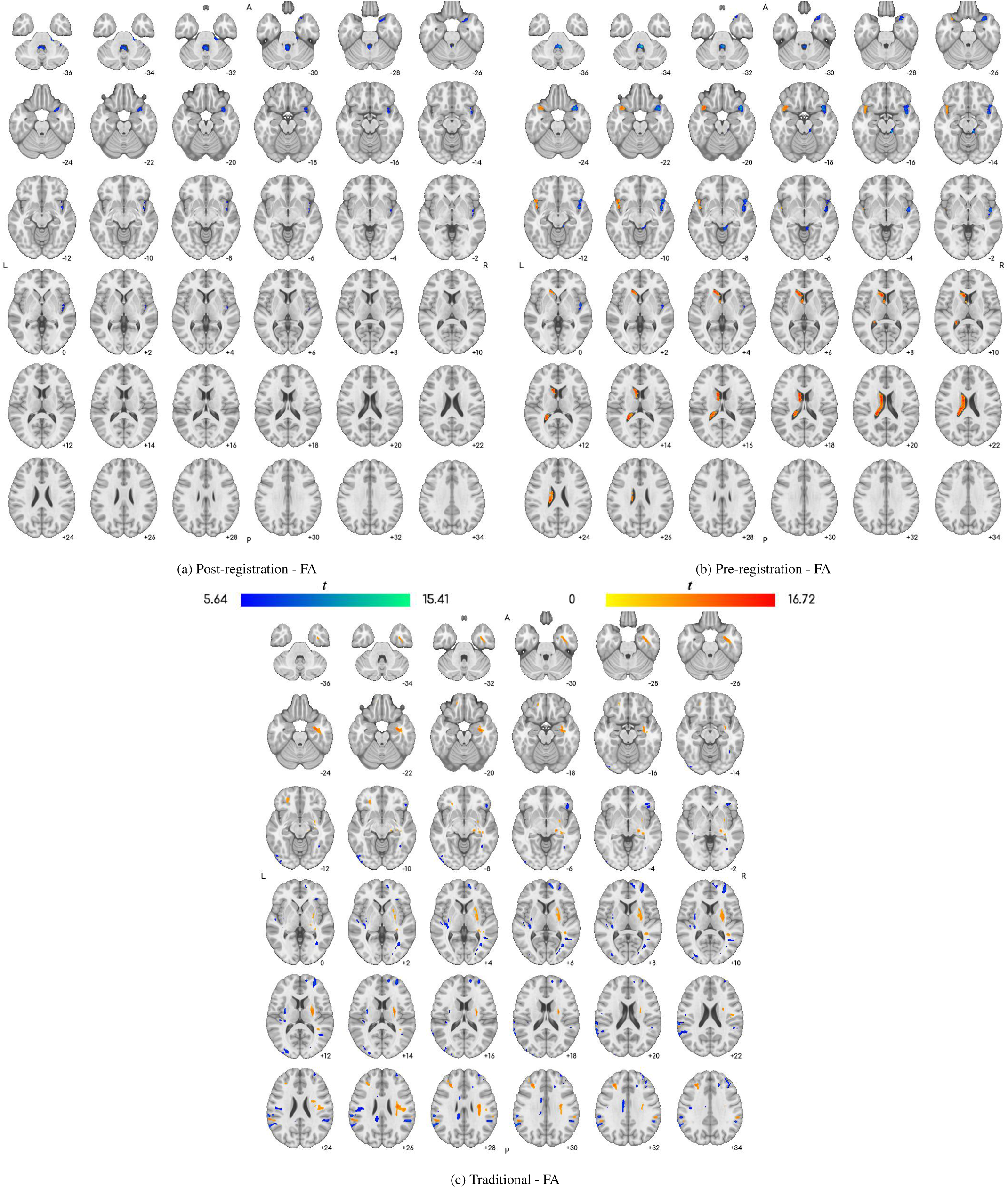
Voxel-based diktiometry (post-registration and pre-registration in the first and second columns, respectively) sensitivity results and traditional voxel-based diffusion analysis results (third column) from *z* = −36 to *z* = +34 in MNI space for the fractional anistropy (FA). Left-dominant PD subjects have been laterally flipped. (Note that patient-right is shown on image-right, i.e. neurological convention.)

For the MD shown in Fig. 4, both CNN based approaches identified a decrease (blue to cyan) in diffusivity generally in the cerebellar white matter and the lenticular nucleus (composed of putamen and pallidum) (lateral to symptom onset). We can also observe an increase (yellow to red) in the diffusivity in the fourth ventricle (not lateralised) and the lateral fissure (lateral to symptom onset) which are likely to result from morphological changes. In the pre-registration version, several structures appeared on both sides of the brain, although with different signs, indicating an asymmetry. One of these structures appears to be the cortico-spinal tract, indicating a laterised effect that is easiest for the network to detect via an asymmetry check. In addition to these asymmetries, the pre-registration approach also found a decrease in diffusivity in the white matter of the temporal lobe.

In the traditional approach, a more slight decrease is seen in the lenticular nucleus. This then extends in the superior direction until the white matter of the cortico-spinal tract. The traditional approach also shows numerous small clusters in cortical regions, especially grey matter regions on the boundary of the external CSF. Due to their small size and distribution, these are likely to be statistical artefacts arising from large patient variability in these regions. Thus, the methods largely agree in the cerebral regions, although the proposed method generally for cerebellar structures to be more indicative of PD.

For the anisotropy, Fig. 5, many of the same regions are identified, although with different signs. For the cerebellum, there is a positive sensitivity to the anisotropy in the caudal region, whereas there is a negative sensitivity in the rostral region closer to the cerebellar grey matter. (This is lateralised for the post-registration approach, but bilateral for the pre-registration approach.) There is also a positive sensitivity to anisotropy in the area surrounding the putamen. In the pre-registration approach, there is a bilateral sensitivity in the cortico-spinal tract adjacent to the thalamus, but again with different signs on the two sides, indicating an asymmetry. One interesting finding is a negative sensitivity in the contralateral ventricle which is present in both CNN types. For the traditional approach, the results using the anisotropy appeared to be similar to those for the fractional anisotropy and will be discussed in the following paragraph.

For the fractional anisotropy, Fig. 6, the traditional VBA approach showed an increase in FA in the cortico-spinal tract (lateral to symptom onset) and a decrease in FA in the putamen. There also appears to be a decrease in the FA in the area of the optical radiations, although that is more difficult to interpret in the context of PD and thus may be artifactual. Interpreting the maps for the CNNs is somewhat more difficult as the sensitivity with respect to FA is derived from those of MD and A as shown in Eq. 4. In the post-registration image, only the fourth ventricle and lateral fissure were identified, suggesting a morphological change. In the pre-registration image, the lateral fissure showed an asymmetry and the contralateral ventricle was also identified, suggesting again a morphological origin.

The pseudo-planarity did not yield any clear results, possibly due to being the smallest source of variation in the diffusion tensor and only well-defined when the other metrics take on high values.

## 5. Discussion

One interesting theoretical difference between the proposed diktiometric method and the traditional voxel-based approach is that the former is sensitive to *non-local patterns* in the underlying voxel values rather than the values themselves. This is crucial in that it allows for the identification of discriminative features conditioned on the entire image even if the marginal distribution of said feature at said voxel is not highly discriminative in itself. The traditional method displays results highly consistent with those of Xiao et al. (2021), despite using a different registration and a different statistical approach, confirming the robustness of the VBA method.

### Classification accuracy

Despite the numerous studies regarding the effects of Parkinson’s disease on white matter and diffusion characteristics in the brain, Prasuhn *et al*. (2020) expressed doubt that DTI could be used for Parkinson’s disease classification after their registration+SVM approach for particular subcortical structures, this approach having some success in classifying other neurological disorders from other data modalities, such as epilepsy diagnosis based on VBM (Chen et al., 2020).) The successful classification between PD patients and HC by these CNNs indicates that the additional flexibility yielded by convolutional neural networks is important to Parkinson’s disease classification and, relatedly, that the descriptive diffusion characteristics of Parkinson’s disease are not localised to a single voxel. Non-linear methods such as a combination of multi-modal MRI features (Talai et al., 2021) have yielded similar accuracies as our CNN approaches, although the approach Talai et al. (2021) used for selecting the “optimal” method involved a degree of data leakage and thus they potentially overestimated their performance on their much smaller dataset.

### Interpreting diktiographic sensitivity maps

Due to the more complex nature of the classification and sensitivity analysis algorithms used, some attention needs to be paid to how these maps are interpreted, specfically cognisant of each other. This is because the pre- and post-registration approaches share many of the same properties, differing only whether or not registration is used to deformably align the input data prior to be inputted into the CNN.

Specifically, this alignment means that morphological information is, by definition, removed as all the images in the preregistered approach are morphologically equivalent to the MNI atlas. Thus, any signal arising from the post-registration approach that does not appear in the pre-registration approach is likely due to morphological changes in the anatomy rather than changes in their diffusion characteristics. This emphasises the necessity to compare the pre- and post-registration approaches during the interpretation of the results especially as some biomarkers (for example, the MD sensitivity in the basal ganglia in Figure 4(a) and (b)) where lateralised and asymmetry features both played a role.

### CNN sensitivity at specific regions

Using the traditional method, we have largely reproduced the results generated by the voxel-based analysis performed by Xiao *et al*. (2021), specifically in terms of the white matter bundle extending from the cortex to the brain stem on the collateral to symptom onset. This validates that the statistical mapping method used for all three approaches is coherent with the literature. (Note that Xiao *et al*. (2021) used a white-matter mask in their approach whereas we did not, leading to a number of grey-matter and sulcal regions to also be identified.) This also validates that simple CNNs trained on a PD diagnosis task do look at relevant regions of the brain.

Notably, our approach appears to generate a number of additional results in comparison to the traditional method. This may confirm Prasuhn *et al*. ‘s (2020) observation that linear classification methods are insufficient. The fact that the traditional method and the pre-registration method rarely overlap also confirms this, suggesting that the regions identified by traditonal VBA are not salient enough for extraction as meaningful features. One clear example of this is the reliance of the neural networks on a mix of cerebellar and cerebral structures, unlike the traditional approach which never found any local biomarkers in the cerebellum, which is in agreement with other recent VBD investigations (Atkinson-Clement et al., 2017). This is of particular interest as the community has been recently calling for more investigation of the cerebellum in the etiology of PD (Mirdamadi, 2016).

Our population-wise results of the post-registration networks show a distinct focus of the neural networks on the basal ganglia and cerebellum, indicating that the network can both identify these regions easily as well as use them to distinguish between PD patients and healthy controls. For the pre-registration network, these locations are also highlighted, indicating that the diffusion characteristics of these regions, rather than their morphology which is removed in the pre-registration workflow, are predictive of PD. What is of particular interest is that these locations are not highlighted in the traditional approach, indicating that the predictive characteristics are truly non-local, referring to a collection of correlated diffusion changes rather than a cluster of pixels that happen to co-vary. The traditional approach appeared to highlight many areas that are not associated with the symptomatology of Parkinson’s disease, likely due to statistical artefacts, whereas the proposed methods highlight areas that are either already known in PD physiopathology (e.g. the basal ganglia) or are under investigation such as the cerebellum.

Our studies agrees with the more general observation made by Xiao et al.Xiao et al. (2021) in that differences between PD patients and controls seems to be lateralised. However, there appear to be a difference in how this lateralisation can be interpreted, specifically between the pre- and post-registration approaches. One possible explanation for this is that the preregistration approach is comparing the two hemispheres, looking for asymmetries which would not be as easily performed by the post-registration network that cannot rely on particular pixels location always representing the same anatomy.

### Role of different diffusion characteristics

Unsurprisingly, MD played the strongest role in the analysis, generating the highest number of significant voxels for both the proposed diktiometric and the traditional voxel-based diffusion analysis approaches. Given that other voxel-based diffusion studies uniformly use MD in their analysis, this is coherent with the literature and confirms the role of MD in understanding the diffusion characteristics of neurological disorders (Atkinson-Clement et al., 2017).

Interestingly, FA appears to have played only a secondary role in the analysis of the CNNs, largely due to its strong connection to MD in its normalisation term. This means that the regions identified by FA sensitivity maps were almost always a subset of those identified by an MD sensitivity, only with a sign inversion as increasing the MD naturally lowered the FA. Although this theoretically could have been overcome by the contribution of the anisotropy (σ) term, this was not observed. Thus, in this sensitivity analysis, it was more meaningful to look to the unnormalised anisotropy term rather than FA to see information traditionally associated with organised microstructure. In addition, we observed the same qualitative results in the traditional voxel-based diffusion analysis. This calls into question the role of FA in diffusion analysis studies more generally as it may be possible that correlated MD increases with FA decreases (and vice versa) are possibly solely due to MD and the contribution of the denominator term, although much more investigation would need to be done to determine what diffusion parameters are sufficiently decoupled to avoid this issue altogether.

The pseudo-planarity appeared to play a very minor role in the analysis, which indicates that the majority of the sensitivity was assigned to the orthogonal MD and A terms. This is unsurprisingly as in the literature, measurements of MD and FA tend to be more significant that the other metrics that measure the shape of the tensor (Abe et al., 2010).

#### 5.1. Limitations and future work Technical limitations of simple CNNs

One of the major technical limitations of this framework involves the *translation invariance* property of traditional, simple CNNs. Although translation invariance is often seen as a strength of CNNs, in the context of medicine, it means that the network has to first be able to localise particular elements before being able to use their diffusion characteristics. This is in contrast to simpler methods such as registration + SVM (Chen et al., 2020) in which the metrics can be extracted from consistent anatomical locations directly as input. This is particularly evident in the post-registration network lacking symmetric features that are opposite in magnitude, indicating an assessment of asymmetry.

However, it should be noted that this limits the degree to which one can use geometric data augmentation techniques, which we hypothesise is one of the reasons why applying CNNs on unregistered images (i.e. the post-registration approach) had a slighter higher performance than when applied on registered ones (i.e. the pre-registration approach). This experiment also relied on a traditional CNN architecture which also leaves open the possibility that a more complex architecture could incorporate more localisation information.

### Methodological limitations

Another theoretical limitation of the study is that multiple CNNs are used. There is a distinct possibility that these networks detect and are sensitive to different non-local diffusion biomarkers. In this case, the sensitivity maps would therefore only indicate features that are commonly and robustly selected by an ensemble of neural networks, with other less robust features remaining invisible.

There is also a possibility that the analysis couples together non-local biomarkers that should be considered independent of each other. Note that there are two ways that a collection of biomarkers may be dependant on each other: they may be correlated in the input data distribution and/or they may be nonlinearly coupled together by the classifier. To give an example of the latter, if the sensitivity maps highlight two regions, there is a possibility that they are coupled together, forming two complementary facets of a singular, more non-local biomarker (for example, an asymmetry biomarker). For traditional VBA, this is not possible as the classifiers themselves are computed independently of each other and thus ensure that the identified features are also treated independent; any coupling between features must be a product of the input data distribution, not the classifier. This “biomarker classification coupling” problem can become even more problematic as we consider multiple nonlocal biomarkers that might interact with each other. We have some evidence that this may be the case, for example in Figure 4 (a) and (b), we see a lateralised biomarker in the basal ganglia (slides -10 to -4) in the post-registration approach, which is bilateral in the pre-registration approach, indicating that the post-registration method is looking solely at the disease onset side whereas the pre-registration approach is looking at the asymmetry between the two sides. This could indicate two separate biomarkers or two different methods for extracting a singular biomarker. In addition, many areas that were highlighted using a single metric (for example, MD) were also highlighted in others, (for example, A and FA) which again could lead to the interpretation of distinct biomarkers or a different facets of a singular biomarker.

In this paper, we heuristically assumed that the different regions of the brain, if highlighted in the sensitivity maps, should be interpreted as independent with the exception of asymmetry checks (i.e. when the same region is highlighted on both sides of the brain with opposite sensitivity signs). We also made the assumption that the metrics were independent of each other. However, an area of future work could be to investigate methods for decoupling the sensitivity maps into components that can vary independently of each other.

### Diagnostic performance and use

Lastly, the classification performance of these networks needs to be improved before they can be used as a diagnostic tool on their own. Although the motivation behind this study was to develop a tool similar to VBA, the end goal of these systems should be to improve the diagnosis of patients more accurately and at earlier time-points in the disease’s progression. Although the methods 70% accuracy is much lower than we have come to expect from deep learning methods in general, it must be noted that it is still, to the best of the author’s knowledge, one of the best in the literature that uses only diffusion tensor information. Not even the T1 structural images, which are often acquired at the same time, are used in the current framework in order for the networks to focus on specifically diffusion-related biomarkers. Thus, there are fours ways we envision this technique to be more useful as a diagnostic tool:

1. the inclusion of more imaging modalities,
2. architectural improvements,
3. prediction of symptom severity, rather than diagnosis,
4. increasing the size of the training dataset, or
5. investigation of a more heterogeneous cohort including early-stage PD.

With the exception of including T1 imaging, this would require extensive research and data collection.

By focusing on symptomatology rather than diagnosis, it may be possible to use these methods in a broader array of clinical contexts related to PD. For example, dementia and other severe cognitive disorders are common counter-indications for deep brain stimulation due to its potentially deleterious impact on cognition, so if cognition-specific biomarkers can be identified, these patients could be more readily screened using imaging (Rodriguez-Oroz et al., 2005). Similarly, more patient-specific maps could be used to help guide deep brain stimulation procedures by targeting areas that are more affected by that patient’s specific symptomatology. In addition, separating the maps based on symptomatology may also help to disentangle the effects specific to PD from those caused by other age-related disorders common in PD cohorts.

### Other areas of future work

Aside from technical development and network optimisation, there is a large potential for the use of more complex diffusion models than the standard diffusion tensor which could still be used in the framework of CNN sensitivity analysis. Specific examples of these models which are already know to be useful in the context of PD include higher order tensors such as those extracted in diffusion kurtosis imaging (Wang et al., 2011) and fibre orientation and density distributions (Xiao et al., 2021). This would also have the benefit of nuancing the results from a scientific perspective, giving us more insight into the diffusion-related effects of PD.

Even within the tensor model, there are still avenues to explore. By acting directly on the diffusion tensors themselves, sensitivity information may be extractable for other information, such as tensor orientation, that is not usually investigated. However, more research would need to be done to ensure that the sensitivity maps for tensor orientation can be meaningfully aggregated (that is, meaningfully combined in a population-wise manner) and interpreted.

## 6. Conclusions

This paper presents *voxel-based diktiometry*, a technique for combining traditional voxel-based analysis with convolutional neural networks in order to identify, visualise, and analyse regions of the brain associated with a particular disorder. The strength of this method comes from the inversion of the traditional VBA paradigm. In VBA, regions of the image that show statistically robust differences are identified which could then be used to help guide diagnosis. In VBD, a strong classifier is used first, and the classifier sensitivity is evaluated for statistical significance, displaying what information is empirically more useful for said classifier. This inversion allows for the regions of interest to display a more non-local character unlike traditional VBA in which (setting aside smoothing and spatial correction) the emphasis is on singular voxels at particular registered locations.

This method has shown evidence for diffusion biomarkers for Parkinson’s disease that are specifically non-local in nature. For example, there is evidence to suggest that assymmetry in the diffusivity of the white matter may be a usable biomarker for lateralisation. These biomarkers are inherently non-local in that they describe how diffusion characteristics in different, separate regions of the brain co-vary and correlate with each other and not only the disease status.

Interestingly, for traditionally convolultional neural-network based classification, it appears that the white matter areas in the cerebellum and brain stem are particularly indicative of Parkinson’s disease, more so than the cerebrum which provides additional evidence of the cerebellar role in PD Mirdamadi (2016).

Overall, voxel-based diktiometry provides an interesting new tool for the investigation of non-local imaging biomarkers of neurological disorders.

## Data Availability

All data was from the Parkinson's Progression Markers Initiative (PPMI) from the Michael J. Fox Foundation.

## Acknowledgements

Alfonso Estudillo Romero is supported through the Fondation Recherche Médicale (FRM) DIC20161236441. John S.H. Baxter is supported by the Institut des Neurosciences Cliniques de Rennes (INCR).

## Data Availability

All data used in this study was taken from the Parkinson’s Progression Markers Initiative (PPMI) created by the Micheal J. Fox Foundation. This data is openly available to the public and no closed data was used in this article.

## Appendix A. Gradient propagation to diffusion metrics

The result of gradient propagation through the network to the input yields the derivatives 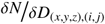 where *D*_(*x,y,z*),(*i, j*)_ is the value of the *i*th row, *j*th column element of the diffusion tensor at location *x, y, z*. As the following is equivalent for all voxels, we will remove *x, y, z* and drop the indexing brackets for the purposes of notation. (Also for notation, both the different and gradient notations are used depending on which is more succinct. For total clarity, ∇_*x*_*y* is a vector whose entries are all *δy*/*δz* where *z* is some element of *x*, which is itself a list or a vector of variables.)

In order to retrieve the values of *μ* and *σ*, it is necessary to perform an eigendecomposition of the diffusion matrix:

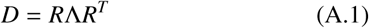

where *R* is an orthonormal matrix and Λ is a non-negative diagonal matrix containing the eigenvalues of the diffusion matrix. Note that this decomposition is not fully unique: it is possible to re-order the elements of Λ and still have a valid matrix. The benefit of *μ* and *σ* is that they are invariant to this ordering whereas other diffusion metrics largely depend on having a specific eigenvalue ordering (e.g. comparing the relative sizes of the largest and second-largest eigenvalues, for example).

Given that *E* is a diagnonal matrix, we can easily derive a simple formula for the elements of *D* in terms of the rotation matrix and eigenvalues, *λ*_*k*_, for some ordering:

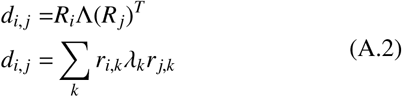

which yields the derivative:

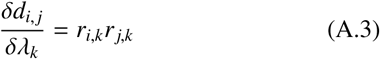

We can then analytically propagate gradients back through this operation under the assumption that *R* is constant with respect to *λ* using the chain rule:

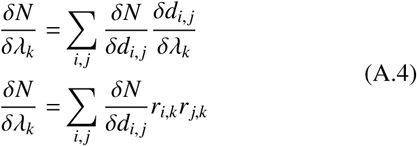

In order to retrieve the gradients with respect to *μ* and *σ*, we propose the following invertible co-ordinate transformation:

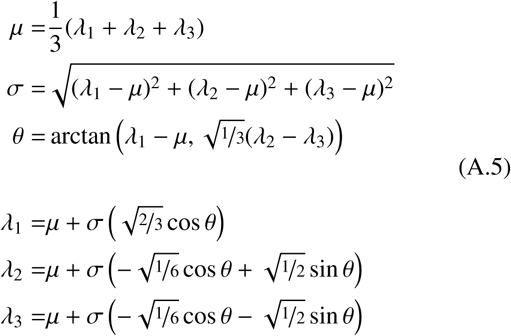

which, using the chain rule, yields the derivatives:

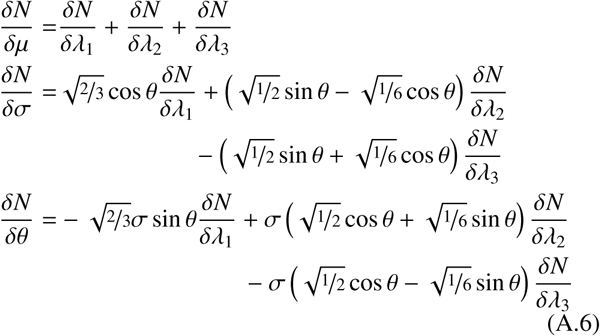

An alternative way of computing the gradients is to consider the above co-ordinate transform as a local basis transformation. The benefit of this transformation is that it is a locally orthogonal basis given *σ* ≠ 0, and thus have orthogonal sensitivities. That is:

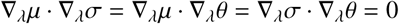

In order to get an orthonormal basis, we have to ensure that the vectors are all unit length, that is, 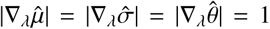. Note that this is already the case for *σ*, i.e. 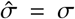 be achieved for *μ* using simple scaling, i.e. 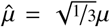, so the last vector 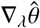 can be found using the cross product: 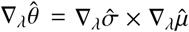. This gives us a magnitude-preserving expression for the sensitivity of the network:

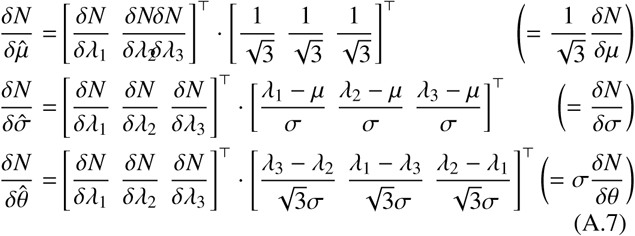

Unlike *μ* and *σ* which are invariant to the ordering of the eigenvalues, *θ* has an ordering dependency, meaning that a consistent ordering must be used for *θ* to be aggregated across a population and *θ* must also be provably continuous at boundary cases where this ordering may be effected by random noise. In the case where the eigenvalues are ascending (i.e. *λ*_1_ ≥ *λ*_2_ ≥ *λ*_3_), *θ* can be thought of as the *pseudo-planarity*, that is an approximate measure of planarity, which describes how much of the remaining degree of freedom is used to make two of the three eigenvalues similar. *θ* = 0 implies that the tensor is as close to being linear (i.e. *λ*_2_ ≈ *λ*_3_) as possible given its mean diffusivity and anisotropy. As *θ* → *π*/3, the tensor is as planar as possible with *λ*_1_ ≈ *λ*_2_ again given the mean diffusivity and anisotropy. As suggested by the trigonometric functions, *θ* also has meaningful units, radians.

Interestingly, the sensitivity with respect to *θ* which is neither cyclic nor has an ordering dependence. This means that our method can measure in a population-wise manner, the sensitivity of the classification network with respect to the shape of the tensor not reflected by its overall size (i.e. mean diffusivity) or overall anisotropy, but by a third, completely orthogonal measure.

